# Environmental drivers of SARS-CoV-2 lineage B.1.1.7 transmission intensity

**DOI:** 10.1101/2021.03.09.21253242

**Authors:** Thomas P. Smith, Ilaria Dorigatti, Swapnil Mishra, Erik Volz, Patrick G. T. Walker, Manon Ragonnet-Cronin, Michael Tristem, William D. Pearse

## Abstract

Previous work has shown that environment affects SARS-CoV-2 transmission, but it is unclear whether emerging strains show similar responses. Here we show that, like other SARS-CoV-2 strains, lineage B.1.1.7 spread with greater transmission in colder and more densely populated parts of England. However, we also find evidence of B.1.1.7 having a transmission advantage at warmer temperatures compared to other strains. This implies that spring and summer conditions are unlikely to slow B.1.1.7’s invasion in Europe and across the Northern hemisphere - an important consideration for public health interventions.

## Introduction

A year into the COVID-19 pandemic, quantifying the factors driving SARS-CoV-2 transmission is still key for the optimal implementation of control strategies. SARS-CoV-2 transmission is influenced by local environmental conditions (Poirier et al., 2020), and we have previously shown that colder temperatures and higher population densities explain spatial and temporal variation in transmission intensity (Smith et al., 2020). A critical insight from previous work has been that human behaviour drives SARS-CoV-2 transmission, and that environment only plays a marginal effect when effective non-pharmaceutical interventions (NPIs) are in place (Poirier et al., 2020, Smith et al., 2020). While several studies have investigated the impact of environmental factors on SARS-CoV-2 transmission during the early stages of the pandemic, numerous new variants have recently emerged (in, for example, the UK; Rambaut et al. 2020, Brazil; Voloch et al. 2020, and South Africa; Tegally et al. 2020) with greater transmissibility than strains present during the early stages of the pandemic (Faria et al., 2021, Volz et al., 2021). Any effect of environment must therefore be re-assessed in the context of a landscape of emerging viral lineages.

The UK SARS-CoV-2 ‘Variant of Concern’ (VOC; also known as lineage B.1.1.7; Rambaut et al., 2020) rapidly spread through the UK population in late 2020, showing greater transmission intensity (as measured by the reproduction number, *R*) than other circulating strains (Volz et al., 2021). The wealth of data collected for this strain makes the VOC a strong test-case for investigating environmental effects in the context of emerging lineages. To assess the impact of environment on VOC transmission, we extracted weekly *R* estimates calculated for fine-grained regions of England (Lower Tier Local Authorities— LTLAs; Mishra et al., 2020), and regressed these against VOC frequency, temperature, and population density for each LTLA. We assessed the impact of the environment by investigating the association between weekly *R* estimates and the predictors (i.e., spatial variation in *R*), for sequential weeks. To account for the VOC’s emergence in the South-East (which is among the warmest and most densely populated parts of England), and its subsequent spread into colder, less densely populated parts of the country, we incorporated interactions between VOC frequency and each of temperature and population density. Without including this interaction, any effect of temperature or population density on *R* might be compounded or masked by spatial autocorrelation with VOC frequency. Following Volz et al. (2021), we measured the effects of the weekly predictors on *R* in the subsequent week, to account for the generation time of SARS-CoV-2.

## Results and Discussion

As observed by Volz et al. (2021), we find a strong positive effect of VOC frequency on transmission intensity, particularly during the initial emerging phase when VOC frequency was less consistent among regions (Table 1). While the UK’s national lockdown initially decreased SARS-CoV-2 transmission intensity, as the VOC spread, *R* increased across regions (Figure 1). We find that the impact of the environment on transmission is mediated by VOC frequency, and it is only at higher VOC frequencies that the effects of temperature and population density are pronounced (Table 2 and Figure 1). Furthermore, we find a greater environmental effect after the UK moved from full lockdown to a (less strict) tiered system, confirming our previous observations that the effect of temperature becomes pronounced when NPIs are relaxed (Smith et al., 2020). Thus, the effects of environmental drivers, and even those of the increased transmissibility of the VOC, are secondary to differences in human behaviour driven by differences in NPIs. As the VOC has spread to dominate the country’s viral population, we now expect environmental factors to become the dominant driver of spatial variation in *R*, with colder areas facing higher transmission intensities than warmer regions, unless NPIs or the accumulation of immunity (either naturally acquired or vaccine derived) sufficiently reduce transmission.

**Table 1:**
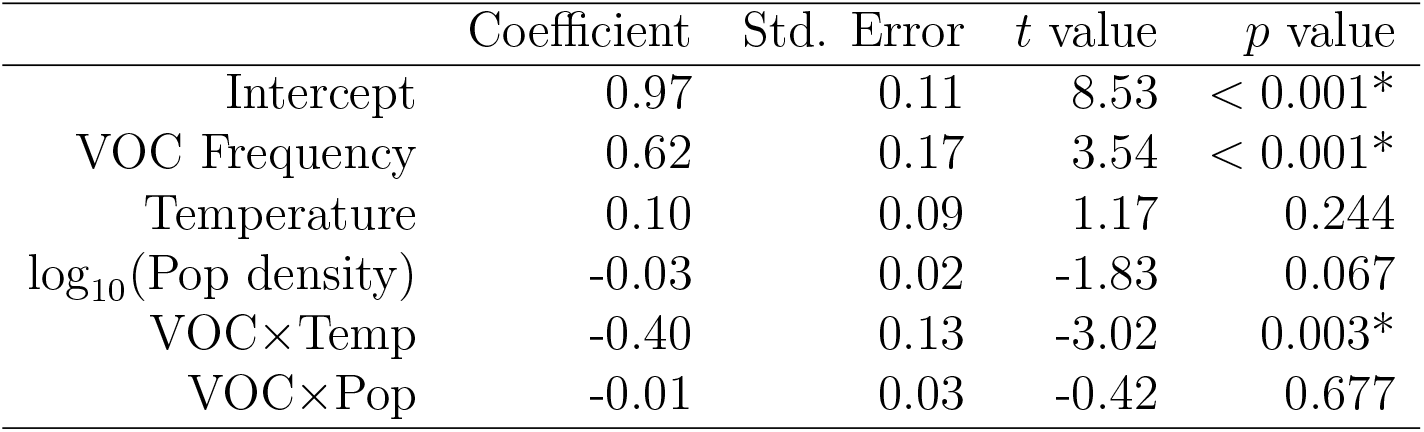
VOC frequency is the strongest driver of transmission during early stages of the VOC sweep (week 46, starting 9 November 2020). Linear regression model with VOC frequency, temperature, and population density as predictors of *R*_*t*_, *r*^2^ = 48%, *F*_5,293_ = 53.5, *p* < 0.001. Predictors were scaled to have mean = 0 and SD = 1 and thus coefficients are measures of the relative importance of each variable. Frequency of the variant is a comparably strong driver of *R*. * = *p* < 0.05.

**Figure 1:**
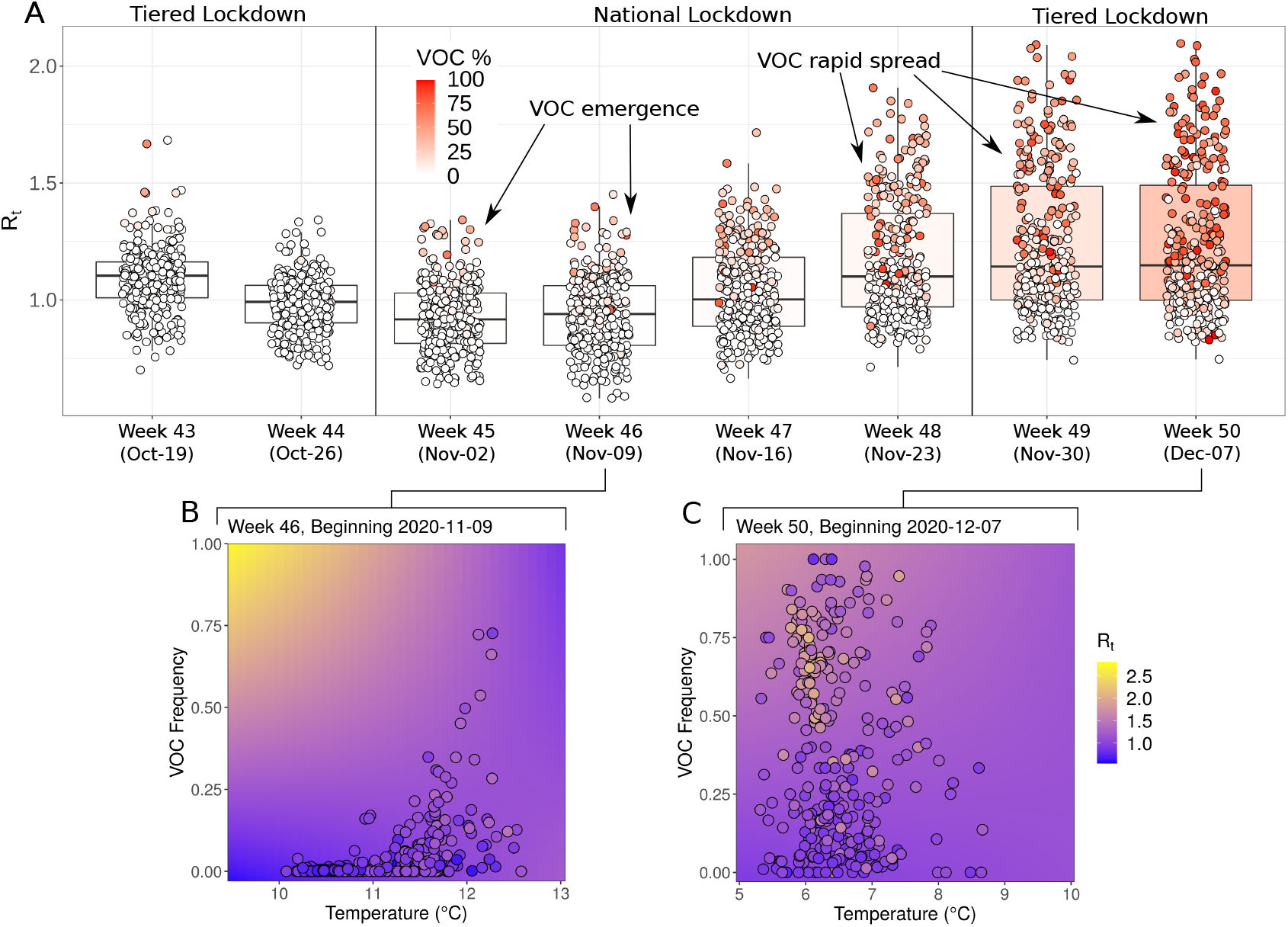
Transmission dynamics during emergence and spread of SARS-CoV-2 lineage B.1.1.7. **A**. Median (bold) and interquartile range (box) of the transmission intensity estimates (*R*) for weeks 43-50 of 2020, with point estimates for each LTLA overlaid. Points are coloured by the frequency of the VOC in positive COVID-19 tests (%), boxplots are coloured by the median VOC frequency across all LTLAs. The first date of each week is given within parentheses. As the VOC emerged, the UK entered a national lockdown, leading to an initial decrease in R across regions. However, as the VOC spread through space to become the dominant lineage, R increased across regions despite the national lockdown. When the UK re-entered a tiered NPI system on the 2nd of December 2020, average *R* further increased across regions. Panels **B** and **C** show the observed (points) and expected (background) effects of temperature and VOC frequency on R during early (week 46) and late (week 50) phases of the variant sweep, respectively (legend shared across both plots).

**Table 2:**
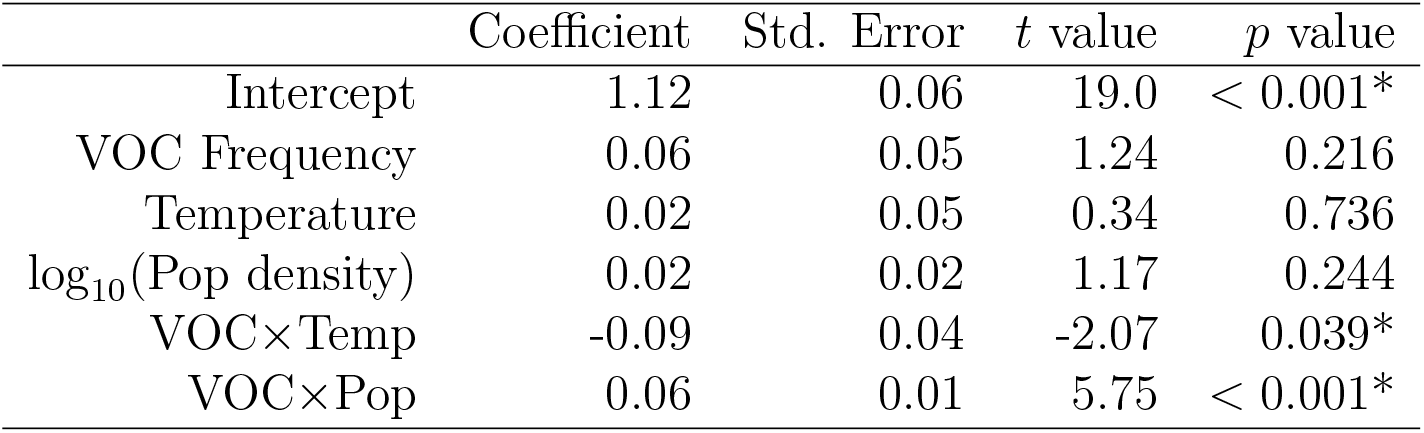
Both temperature and population density significantly interact with VOC frequency during late stages of the VOC sweep (week 50, starting 7 December 2020). *r*^2^ = 55%, *F*_5,298_ = 71.6, *p* < 0.001. Predictors were scaled to have mean = 0 and SD = 1. Significant interaction effects between variant frequency and environmental parameters are observed. Significant interactions show that at high VOC frequencies, there is a negative effect of temperature and a positive effect of population density on *R*_*t*_. * = *p* < 0.01.

| | Coefficient | Std. Error | $t$ value | $p$ value |
| --- | --- | --- | --- | --- |
| Intercept | 1.12 | 0.06 | 19.0 | < 0.001* |
| VOC Frequency | 0.06 | 0.05 | 1.24 | 0.216 |
| Temperature | 0.02 | 0.05 | 0.34 | 0.736 |
| $\log_{10}(\text{Pop density})$ | 0.02 | 0.02 | 1.17 | 0.244 |
| VOC×Temp | -0.09 | 0.04 | -2.07 | 0.039* |
| VOC×Pop | 0.06 | 0.01 | 5.75 | < 0.001* |

To investigate the potential role of immunity in our results, we perform a sensitivity analysis and re-run our regression analyses on *R* estimates corrected for the relative attack rate (AR) estimates. This accounts for the potential slowdown in *R* due to the accumulation of natural immunity across the population through time. We obtain qualitatively identical results in the AR-corrected analyses, but generally lower correlation (additive decrease in of 16% in weeks 45 and 46) as the VOC emerged, versus in the later stages (average decrease in of only 3%, Supplementary Table S1). Note that the time-frame of our analyses was prior to mass vaccination of the UK public and even in the latest time-point (week 50), median attack rate across regions was less than 10% (Supplementary Figure S1).

We additionally investigated whether the VOC may respond differently to the environment as compared to non-VOC strains. Volz et al. (2021) estimated the transmission intensity of VOC vs non-VOC strains and here we regressed the ratios of VOC *R* to non-VOC *R* against temperature, to understand under which environmental conditions the VOC may have an advantage. This was performed at a larger spatial scale than our LTLA analyses (English NHS sustainability and transformation partnerships – STP regions). We found a small, positive correlation between temperature and the *R* ratio (Supplementary Table S2), *i*.*e*., at warmer temperatures the VOC was even more transmissible than non-VOC strains, than in colder conditions. This may be expected, as differences in fitness between strains are likely to be magnified in harsher environments where fitness is reduced overall (Soberon and Peterson, 2005). This effect may have played into the dynamics of the spread of the VOC in the South East of England by enhancing its spread even more in regions where previous strains’ transmission intensities were lower.

Whereas, previously, a central question has been whether winter conditions would enhance SARS-CoV-2 transmission, in the context of these new strains it is important to ask whether warmer summer conditions could slow transmission. Here we have shown that temperature can drive variation in transmission intensity between regions and between variants, but this effect is likely secondary to the overall increased transmission intensity of the new variant. Our results show that variant B.1.1.7 is more transmissible in colder areas. Using the model fitted to the week 50 data, our results suggest that when the variant accounts for 100% of SARS-CoV-2 cases in the population, *R* increases by approximately 0.12 per °C of temperature decrease. However, we also find that in warmer conditions, the difference in transmissibility between VOC and non-VOC strains is greater. Our results suggest that for each °C of temperature increase, the ratio of VOC to non-VOC *R* may increase by approximately 0.12-0.22 (Supplementary Table S2).

While we caution against extrapolating from winter to summer conditions, our results highlight that there is no reason to suppose that summer weather alone will slow down the invasion dynamics of B.1.1.7 and significantly reduce the transmission intensity of SARS-CoV-2. Thus, it is imperative to quantify and continue to monitor the impact of interventions (*e*.*g*., vaccines and NPIs) to inform policy. In that regard, the speed with which the VOC has spread through the UK, while concerning from a public health perspective, provides the perfect opportunity to parameterise models of its responses. Importantly in the context of new strains, transmission of respiratory viruses in tropical climates (such as Brazil, where a new variant has arisen; Voloch et al., 2020) are often more sensitive to rainfall than temperature (Pica and Bouvier, 2012). Better characterisation of past, and new potential relationships between environment and transmission, as new variants emerge and spread internationally, is a research priority.

## Materials and Methods

### Epidemiological data collection

Weekly transmission intensity (*R*) estimates for each of England’s lower-tier local authorities (LTLAs) were obtained from a Bayesian epidemiological model (Mishra et al., 2020). The weekly VOC frequencies were calculated for each LTLA as the percentage of true positive rate (TPR)-adjusted S-gene target negative test to positive test results, as in Volz et al. (2021). Estimates of *R* for VOC and non-VOC strains for England’s NHS sustainability and transformation partnerships (STPs) were obtained from Volz et al. (2021).

### Environmental data collection

Population density and daily temperature data were computed for LTLAs and STPs using the same methods as Smith et al. (2020). Briefly, we collected global population density data from the Center for International Earth Science Information Network (CIESIN) (2018), and hourly temperature (*T*) estimates from the Copernicus Climate Change Service (2020) at a 0.25×0.25° spatial resolution. The Climate Data Operators program Schulzweida (2019) was used to to compute daily means from the hourly temperatures, which were subsequently combined into weekly means. We then averaged the population density and temperature values (median) across each spatial unit (LTLAs or STPs) given in shapefiles acquired from the The Open Geography portal from the Office for National Statistics (https://geoportal.statistics.gov.uk/).

### Regression analysis of environmentally driven transmission

We performed multiple linear regression on *R* estimates with VOC frequency, temperature and popluation density as predictors, allowing for interactions between VOC frequency and each of the other predictors, *i*.*e*.:

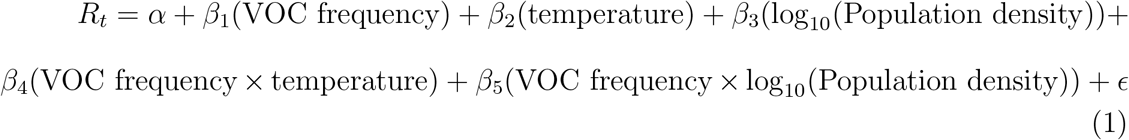

This was performed separately for sequential weeks, with the weekly predictors regressed against *R* estimates for the subsequent week, to account for the generation time of SARS-CoV-2.

### Sensitivity analysis using attack rates

As the virus spreads through the population, so background immunity increases in the population and thus there are fewer potential targets for the virus to infect, ultimately leading to a reduction in the transmission rate. If the effect of immunity is suitably large, this may impact our results. We therefore performed a sensitivity analysis on our regression analyses by correcting the *R* estimates by the attack rate (AR), which is the cumulative infections per population in each area. We calculated corrected-*R*_*t*_ as 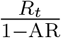 and used this as the outcome variable in our regression models (supplementary information).

### Regression analysis of variant transmission rate

To test whether there were differences in the environmental responses of VOC transmission version non-VOC transmission rates, we regressed the ratio of VOC to non-VOC *R* against temperature. We used two types of models, one with a fixed effect for each area (STP regions) and one with a random effect for area. These models were fit jointly to the data across weeks 45-50 and fixed effects of the epidemiological week were included in both cases (supplementary information).

## Data Availability

No new data was collected for this study. The data used is archived in a github repository alongside the analysis code.

https://github.com/smithtp/covid19-variant-N501Y

## Data and code availability

All code to replicate these analyses and associated data are available from the following github repository: https://github.com/smithtp/covid19-variant-N501Y

## Notes

### Competing Interest Statement

The authors have declared no competing interest.

### Funding Statement

This work was funded by a Natural Environment Research Council grant NE/V009710/1 and by the Imperial College COVID-19 Research Fund.

