## Supplementary information for "Environmental drivers of SARS-CoV-2 lineage B.1.1.7 transmission intensity"

### Supplementary material for: Environmental drivers of SARS-CoV-2 lineage B.1.1.7 transmission intensity

#### Sensitivity analysis accounting for immunity

As the virus spreads through the population, so background immunity increases in the population and thus there are fewer potential targets for the virus to infect, ultimately leading to a reduction in the transmission rate. If the effect of immunity is suitably large, we may not be able to directly compare estimates of $R$ through time, due to this landscape of changing immunity. We can account for this by correcting the estimates of $R$ by the attack rate (AR) of the virus, which is the cumulative infections per population size of a given area. We calculate corrected-$R_t$ as $\frac{R_t}{1-AR}$ and use this as the outcome variable in our regression models. We find lower $r^2$ for the AR-corrected models, particularly in earlier weeks of the VOC spread (table S1). Additionally, when immunity is accounted for, we see stronger negative effects of temperature on $R_t$, but lesser effects of the interaction between temperature and VOC. In most regions, the attack rate is fairly low (median AR $< 10\%$), so it is unsurprising that this has a relatively small effect on our model outcomes (see Figure S1).

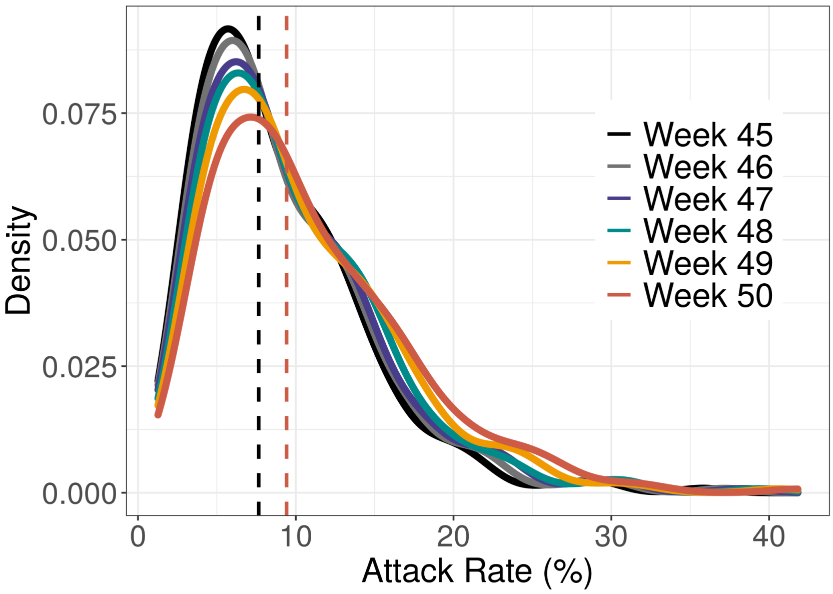


**Figure S1: Distribution of attack rates across LTLAs through time.** Dashed lines show median AR for week 45 (7.65%, black) and week 50 (9.42%, red) respectively, showing a small increase in naturally acquired immunity through time.

**Table S1: Differences in model coefficients and $r^2$ when using the AR-corrected $R_t$ versus the standard $R_t$ as the outcome variable in regression models.** Accounting for immunity generates a marginally worse fit to the data, as shown by decreases in $r^2$.

| Week | ΔIntercept | ΔVOC | ΔTemp | ΔPop | ΔVOC×Temp | ΔVOC×Pop | $\Delta r^2$ |
|---|---|---|---|---|---|---|---|
| 45 | 0.077 | -0.014 | 0.029 | -0.002 | 0.127 | 0.011 | -0.16 |
| 46 | 0.211 | 0.081 | -0.092 | -0.014 | -0.037 | 0.016 | -0.16 |
| 47 | 0.117 | -0.005 | -0.086 | -0.004 | 0.159 | 0.006 | -0.06 |
| 48 | 0.073 | 0.057 | -0.065 | -0.005 | 0.061 | 0.008 | -0.02 |
| 49 | -0.012 | 0.073 | -0.093 | -0.012 | 0.037 | 0.008 | -0.01 |
| 50 | 0.065 | 0.036 | -0.068 | -0.014 | 0.020 | 0.007 | -0.02 |

#### Analysis of $R_t$ ratios

We tested whether the VOC responds differently to temperature than non-VOC strains by regressing the ratios of VOC and non-VOC $R_t$ against temperature. We fit these models jointly to the data across weeks 45-50, examining the role of time (i.e., week) in explaining variance of $R_t$ using fixed effects and examining the role of geography using both fixed and random effects. We find a positive effect of temperature on the ratio of VOC to non-VOC $R_t$ i.e., the VOC is even more transmissible than non-VOC strains when it is warmer (Table S2).

**Table S2: Effect of temperature on ratio of $R_t$ (VOC $R_t$/non-VOC $R_t$).** In models with both fixed and random effects of geography, we see a positive effect of temperature on the ratio of between VOC and non-VOC strains. 95% CIs for the temperature coefficient exclude zero in both models and therefore $p < 0.05$.

| Model | Temperature Coefficient (95% CI) |
|---|---|
| Fixed | 0.22 (0.14 – 0.30) |
| Random | 0.12 (0.04 – 0.20) |
